# Cation Enrichment and Hypersialylation in Chronic Rhinosinusitis Mucus

**DOI:** 10.64898/2026.05.23.26353957

**Authors:** Amanda M. Wood, Rachel E. Detwiler, McKenna Coughlin, Chelsea E. Pollard, Jeremiah A. Alt, Abigail Pulsipher, Jessica Kramer Stratton

**Author notes:** indicates equal contribution (Amanda M. Wood, Rachel E. Detwiler).

## Abstract

**Background:** Chronic rhinosinusitis (CRS) is a heterogeneous inflammatory airway disease associated with impaired mucociliary clearance and persistent inflammation. While prior work has focused on inflammatory and molecular pathways, the physicochemical properties of mucus itself remain poorly characterized. This study aimed to define compositional and biophysical features of CRS mucus that may contribute to dysfunction.

**Methods:** A prospective cross-sectional study was conducted in 15 adults undergoing endoscopic sinus surgery (11 CRS, 4 controls). Mucus was collected from the middle meatus. Hydration was measured by lyophilization. Ionic composition was quantified using mass spectrometry. Viscoelasticity was assessed via oscillatory shear rheology. Total protein, total carbohydrate, sialic acid (Sia) and fucose (Fuc) content were quantified using enzymatic and chemical assays. Statistical comparisons were performed using nonparametric tests.

**Results:** CRS mucus exhibited significantly higher Ca^2+^ and Mg^2+^ concentrations (approximately two-fold; p<0.05) and increased variability in hydration and ion content compared to controls. Rheology showed greater heterogeneity and a non-significant trend toward increased viscoelasticity in CRS. Total protein and carbohydrate content were not significantly different; however, the carbohydrate-to-protein ratio was significantly reduced in CRS (p=0.04). Sia content and Sia-to-carbohydrate ratio were significantly elevated in CRS (p=0.04 and p=0.002), particularly in CRS with nasal polyps. Fuc content did not differ between groups.

**Conclusions:** CRS mucus demonstrates coordinated alterations in ionic composition and glycosylation, characterized by increased cation content, hypersialylation, and reduced carbohydrate-to-protein ratios. These changes may contribute to altered mucus properties and impaired mucociliary clearance, highlighting mucus composition as a potential therapeutic target in CRS.

## 1. Introduction

Chronic rhinosinusitis (CRS) is a heterogeneous inflammatory disease of the upper airways that imposes a substantial burden on quality of life and healthcare systems.^1^ A defining feature of CRS is impaired mucociliary clearance (MCC), which contributes to mucus stasis, persistent inflammation, and susceptibility to microbial colonization.^2–4^ While inflammation and epithelial dysfunction are well-recognized components of CRS pathophysiology, the role of mucus itself, as both a barrier and disease mediator, remains incompletely understood.^5–7^

Airway mucus is a complex hydrogel composed primarily of water, ions, and high-molecular-weight mucin glycoproteins.^8^ Gel physical properties, including hydration, viscoelasticity, and transport behavior, are critical for normal MCC. In other airway diseases, relatively small changes in hydration or composition are known to produce substantial alterations in rheology and clearance.^3,9^ In CRS, mucus abnormalities are commonly observed clinically; however, the underlying biochemical and biophysical drivers remain poorly defined. Prior CRS mucus studies have focused almost exclusively on mucin gene expression or inflammation-related proteomics^10–18^, with limited direct and broader characterization of mucus itself. This gap has limited understanding of how hydration, ionic environment, and molecular composition collectively regulate mucus function in CRS.

Mucin saccharides, particularly (Fuc) and sialic acid (Sia) glycans, contribute to mucus biophysics and biochemistry.^19–23^ Glycans influence mucin conformation, charge, hydration, and polymer network properties affecting rheology, as well as regulate immune and infection processes.^24–27^ Fuc and Sia are reported as altered in multiple respiratory diseases.^28–32^ CRS sialylation and fucosylation have been previously investigated;^33–36^ however, tissue rather than mucus was used in most cases and the methods employed are notorious for poor specificity^37,38^ and nasal polyposis was not considered.^1^

The primary objective of this study was to comprehensively characterize CRS mucus by integrating analyses of hydration, ions, rheology, and biochemical composition, including total protein and carbohydrate content, and specific glycans Fuc and Sia. By comparing CRS to non-CRS controls, we aimed to identify physicochemical features that distinguish disease-associated mucus. This integrated analysis may help link molecular alterations to clinically relevant outcomes, including impaired MCC, microbial burden, and disease severity and persistence.

## 2. Methods

### 2.1 Study participants and data collection

Mucus samples were collected from an ongoing prospective, observational cross-sectional study. Fifteen adults (>18 years) were enrolled after informed consent under a University of Utah Institutional Review Board– approved protocol (IRB no. 00074325). Inclusion criteria comprised patients with symptomatic CRS meeting American Academy of Otolaryngology–Head and Neck Surgery criteria^39^ and whom elected to undergo endoscopic sinus surgery (ESS), while control subjects were defined as patients without CRS electing endoscopic surgery for nasal obstruction. Exclusion criteria included oral corticosteroid use within 2 weeks, prior biologic therapy, recurrent acute rhinosinusitis, cystic fibrosis, granulomatosis with polyangiitis, autoimmune disease, and myelodysplastic syndrome. Demographic and clinical data collected included age, sex, ethnicity, and relevant comorbidities (allergy, asthma, headache, diabetes, gastroesophageal reflux disease, and NSAID-exacerbated respiratory disease). All participants underwent standardized evaluations including history, physical examination, computed tomography (CT), and nasal endoscopy. Disease severity was assessed using Lund–Mackay and Lund–Kennedy scoring systems. Polyp status was recorded intraoperatively, and CRS patients were stratified into CRS with nasal polyps (CRSwNP) and without nasal polyps (CRSsNP) subgroups.

### 2.2 Region selection for sinonasal mucus sampling and collection procedures

The middle meatus was selected for mucus collection, as it is physiologically representative of CRS and serves as the primary drainage pathway for the frontal, maxillary, and anterior ethmoid sinuses. Obstruction or discharge in this region is a key diagnostic feature of CRS.^40^ During ESS, mucus samples were collected from the left and right middle meatus by gentle aspiration into a 50-mL conical tube under vacuum control. The mucus was transferred to microcentrifuge tubes, flash frozen on dry ice, and stored at −80 °C until used as described in sections 2.3–2.8.

### 2.3 Mucus hydration and ion analysis

Mucus water content was analyzed by mass displacement after lyophilization. Three x 20 µL of each mucus sample were weighed on a Cahn C-33 microbalance, frozen at –20 °C and then lyophilized on a Labconco FreeZone 2.5L −84°C benchtop lyophilizer at 0.020 mbar for 24 hours. Dry sample weights were obtained, and the mass fraction of water was calculated by mass loss as the average of the three replicates. Lyophilized samples were digested in concentrated nitric acid (trace metal grade) in a PTFE vial at 150°C, then diluted to 5 mL in ultrapure water. Samples were analyzed by triple quadrupole inductively coupled plasma mass spectrometry (ICP-QQQ) on an Agilent 8900.

### 2.4 Mucus rheology

Rheological measurements were performed on a NETZSCH Kinexus Prime Ultra+ rheometer at 34 ± 0.1 °C using a humidified solvent trap. Mucus samples were thawed on ice, and 27 µL loaded onto a roughened parallel plate (8 mm diameter) and equilibrated for 5 min at 34 °C. Amplitude sweeps (1 Hz, 0.01% strain to the end of the linear viscoelastic regime [LVER]) were used to determine the measurement strain, defined by a sustained increase in phase angle. A strain of 2% (maximum within the LVER) was used for all subsequent measurements. Frequency sweeps were performed from 0.1–20 Hz and back at 2% strain, with 10 points per decade. Data were interpolated (cubic polynomial) to 300 matched frequencies, and forward and reverse sweeps were averaged. Results were exported at 6 points per decade. Storage modulus (G′), loss modulus (G″), complex modulus (G*), and phase angle (δ) are reported at 1 and 10 Hz. Four independent runs were performed per sample.

### 2.5 General mucus preparation and homogenization for biochemical assays

Frozen mucus samples were thawed on ice and serially diluted in ultrapure water to fall within assay sensitivity and calibration ranges. Samples were transferred using a positive displacement pipette and homogenized on ice using a water bath sonicator (0.5–4 h) until visually transparent. Persistent aggregates were disrupted by probe sonication in an ice bath (50% amplitude, 2 s bursts). All assays were performed in clear 96-well plates (Greiner) using a Molecular Devices SpectraMax M2 plate reader. Tube cap clips were used during heating steps to prevent sample loss.

### 2.6 Mucus total protein content

Total protein content was quantified via commercial bicinchoninic acid (BCA) assays (Thermo Scientific) according to manufacturer instructions. A minimum of three aliquots of each homogenized mucus sample were analyzed in triplicate. The detailed procedure can be found in the SI.

### 2.7 Mucus total carbohydrate content

Total carbohydrate content was quantified via commercial phenol-sulfuric acid assay (Cell Biolabs Inc.) according to manufacturer instructions. A minimum of two aliquots of each mucus sample were analyzed in triplicate. The detailed procedure can be found in the SI.

### 2.8 Mucus Sia and Fuc content

Fuc content was quantified using a commercial L-fucose dehydrogenase assay kit (Megazyme) according to manufacturer instructions. Sia content was quantified via an improved Warren Method using a commercial kit (Sigma-Aldrich) according to manufacturer instructions. A minimum of two aliquots of each mucus sample were analyzed in triplicate for each assay and detailed procedures can be found in the SI.

### 2.9 Statistical analyses

Patient mucus samples were analyzed using 2–4 separate mucus aliquots which were independently analyzed in triplicate, or as specifically indicated below. Data were processed using rSpace2.1.0.0, MATLAB R2025b, Graphpad Prism 10, and Microscoft Excel. Non-parametric Mann-Whitney t-tests were utilized to compare controls vs. all CRS samples. Non-parametric ANOVA Kruskal-Wallis tests were used to compare controls vs. endotypes (CRSsNP and CRSwNP) with follow-up multiple comparison tests. Hydration, ions, rheology, protein, and carbohydrate values are reported as the mean ± standard deviation (SD). The coefficient of variance (CV) is also reported to describe spread of different datasets. Significance is defined as *p* < 0.05. A table of all statistical data can be found in Supplementary Information (SI).

## 3. Results

### 3.1 Demographics and clinical characteristics

Fifteen participants were enrolled in this pilot study. Four participants had no CRS diagnosis and served as controls, while 11 had an active CRS diagnosis. Among those with CRS, 6 were male and 5 were female, and all were White. In the control group, 3 participants were male and 1 was female; 3 were White and 1 was of another race. Participant Lund-Mackay CT score, Lund-Kennedy endoscopy score, allergy, asthma, headache, diabetes, GERD, and AERD status are included in Table 1. No significant differences in these variables were observed between cohorts.

**Table 1.** Patient demographics.

|  | Control<br>(n=4) | CRS<br>(n=11) | *p*-<br>value |
| --- | --- | --- | --- |
| **Sex** *N* (%) |  |  | 0.52 |
| **Male** | 3 (75.0%) | 6 (54.5%) |  |
| **Female** | 1 (25.0) | 5 (45.5) |  |
| **Age** (mean ± SD) | 45.0 (22.4) | 65.0 (19.0) | 0.29 |
| **Race\*** *N* (%) |  |  | 0.27 |
| **White** | 3 (75.0) | 11 (100.0) |  |
| **Other** | 1 (26.0) | 0 (0.0) |  |
| **Lund-Mackay CT Score** (mean ± SD) | 3.0 (4.2) | 11.0 (6.5) | 0.17 |
| **Lund-Kennedy Endoscopy Score** (mean ± SD) | 2.7 (3.1) | 4.1 (3.1) | 0.61 |
| **Nasal Polyp Status** *N* (%) | 0 (100.0) | 7 (63.6) | 0.077 |
| **Allergy\*** *N* (%) | 3 (75.0) | 7 (63.6) | >0.99 |
| **Asthma** *N* (%) | 2 (50.0) | 6 (54.5) | >0.99 |
| **Headache** *N* (%) | 0 (0.0) | 0 (0.0) | >0.99 |
| **Diabetes** *N* (%) | 0 (0.0) | 0 (0.0) | >0.99 |
| **GERD** *N* (%) | 1 (25.0) | 3 (27.3) | >0.99 |
| **AERD** *N* (%) | 0 (0.0) | 3 (27.3) | 0.52 |

GERD: gastroesophageal reflux disease; AERD: aspirin-exacerbated respiratory disease; CT: computed tomography; \*subject-reported

### 3.2 Sinonasal mucus water and ion content

Mucus hydration strongly influences MCC and transport of drugs, microbes, and immune components.^2,41–43^ We quantified water content by comparing hydrated and dry masses and found no significant difference between CRS and control groups (Fig. 1A). However, variability was greater in CRS, which ranged from 90.6–98.6% water (CV= 2.59%), compared to 95.1–98.6% (CV= 1.57%) in controls. No differences were observed by polyp status (see SI).

**Figure 1.**
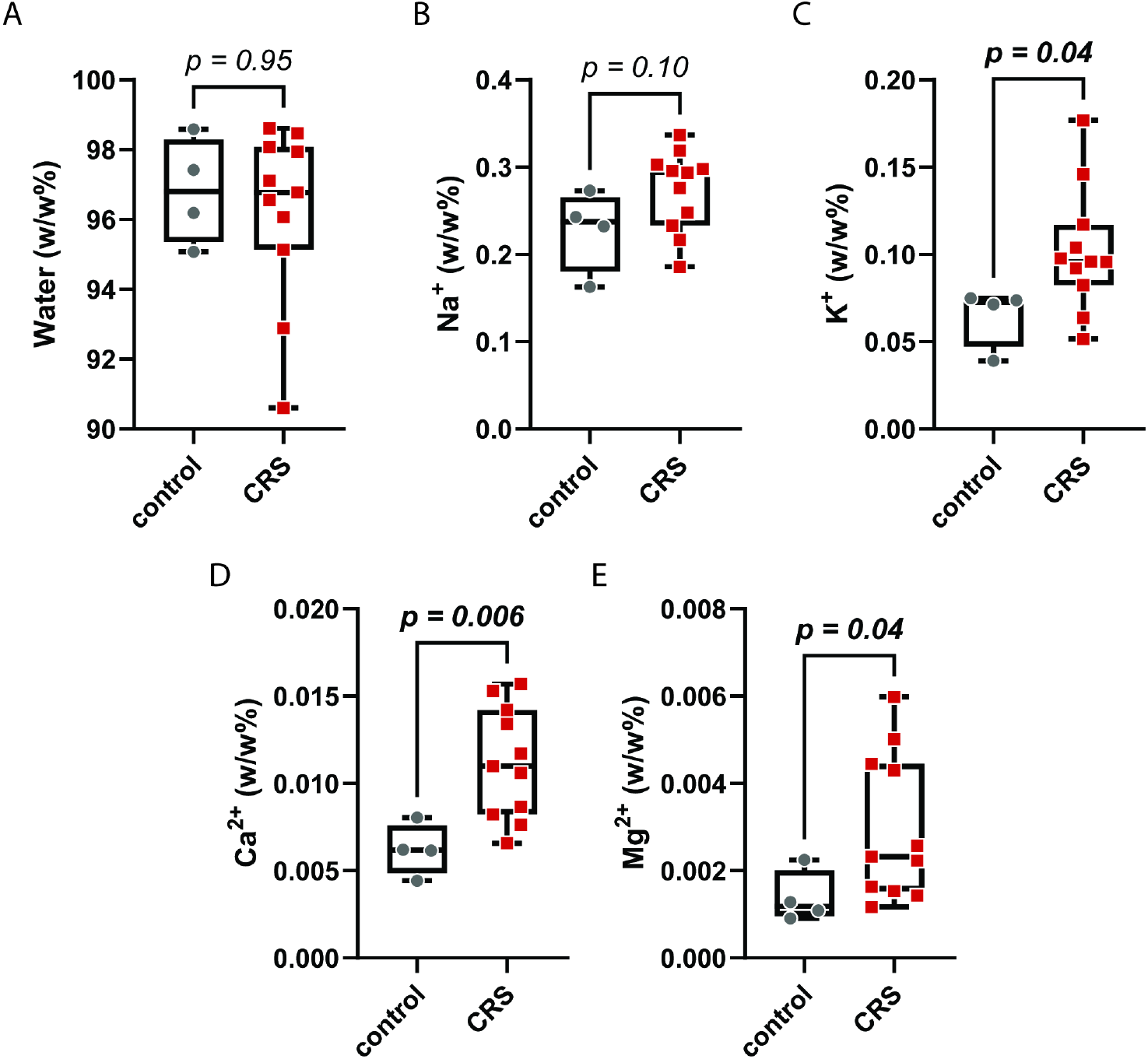
Hydration and cation content (w/w %) of CRS mucus (*n* = 11) and control mucus (*n* =4) where A) is mucus water content by lyophilization mass loss; and B–E) is mucus cation content by ICP-QQQ. Significant increases in C) K^+^ (*p* = 0.04), D) Ca^2+^ (*p* = 0.006) and E) Mg^2+^ (*p* = 0.04) were measured in CRS compared to controls. Mann-Whitney t-tests were performed to determine significance between groups.

Inorganic cations interact with anionic mucin residues (e.g., Glu, Asp, Sia, sulfated glycans), with monovalent ions screening charge and divalent ions promoting cross-linking. ICP-QQQ was used to quantify Na^+^, K^+^, Ca^2+^, and Mg^2+^, reported as w/w% hydrated mass. Monovalent ions dominated, with Na^+^ most abundant, followed by K^+^, Ca^2+^, and Mg^2+^ (Na^+^ ≫ K^+^ ≫ Ca^2+^ > Mg^2+^), a pattern preserved across groups and regardless of polyp status (Fig. 1B–E; SI). Despite similar relative distributions, CRS samples showed greater variability and overall higher ion content by weight.

Specifically, Ca^2+^ and Mg^2+^ were approximately two-fold higher in CRS compared to controls (0.011 ± 0.003 vs. 0.0062 ± 0.001 w/w%, *p* = 0.006; and 0.0030 ± 0.0020 vs. 0.0014 ± 0.0006 w/w%, *p* = 0.04, respectively). K^+^ was modestly increased (CRS = 0.10 ± 0.035 vs. control = 0.065 ± 0.017 w/w%, *p* = 0.04), while Na^+^ trended higher but was not significant (*p* = 0.10). Subgroup analysis showed increased Na^+^ (*p* = 0.04) and Ca^2+^ (*p* = 0.02) in CRSsNP vs. controls, but with no differences between CRS subtypes (see SI).

### 3.3 Sinonasal mucus rheological properties

Mucus hydration and ionic crosslinking strongly influence rheological properties, and thus MCC and transport.^2,41–43^ Given observed differences in water and cation content, we assessed mucus viscoelasticity using oscillatory shear rheology. Storage (G′) and loss (G″) moduli represent elastic and viscous behavior, respectively, while complex modulus (G*) and phase angle (δ) reflect overall stiffness and viscoelastic balance. Measurements at 1 vs. 10 Hz approximate physiologic deformation rates in the airway comparable to breathing vs. ciliary beating, coughs, and sneezes.^44,45^

At 1 Hz, all samples exhibited gel-like behavior (G′ > G″; Fig. 2A–D). Control mucus showed relatively uniform properties (G′ = 12.7 ± 1.8 Pa, CV = 14.3%; G″ = 1.84 ± 0.62 Pa, CV = 33.8%), whereas CRS samples were more variable and mean moduli trended higher (G′ = 17.9 ± 11.9 Pa, CV = 66.4%; G″ = 3.20 ± 3.8 Pa, CV = 118%). Similar trends were observed for G* (control: 12.8 ± 1.9 Pa, CV = 14.6%; CRS: 18.3 ± 12.4 Pa, CV = 67.5%). Phase angle indicated predominantly elastic behavior in both groups, again with greater variability in CRS (control: 7.93 ± 2.4°, CV = 30.1%; CRS: 8.30 ± 4.5°, CV = 54.0%). No differences reached statistical significance (*p* > 0.05).

**Figure 2.**
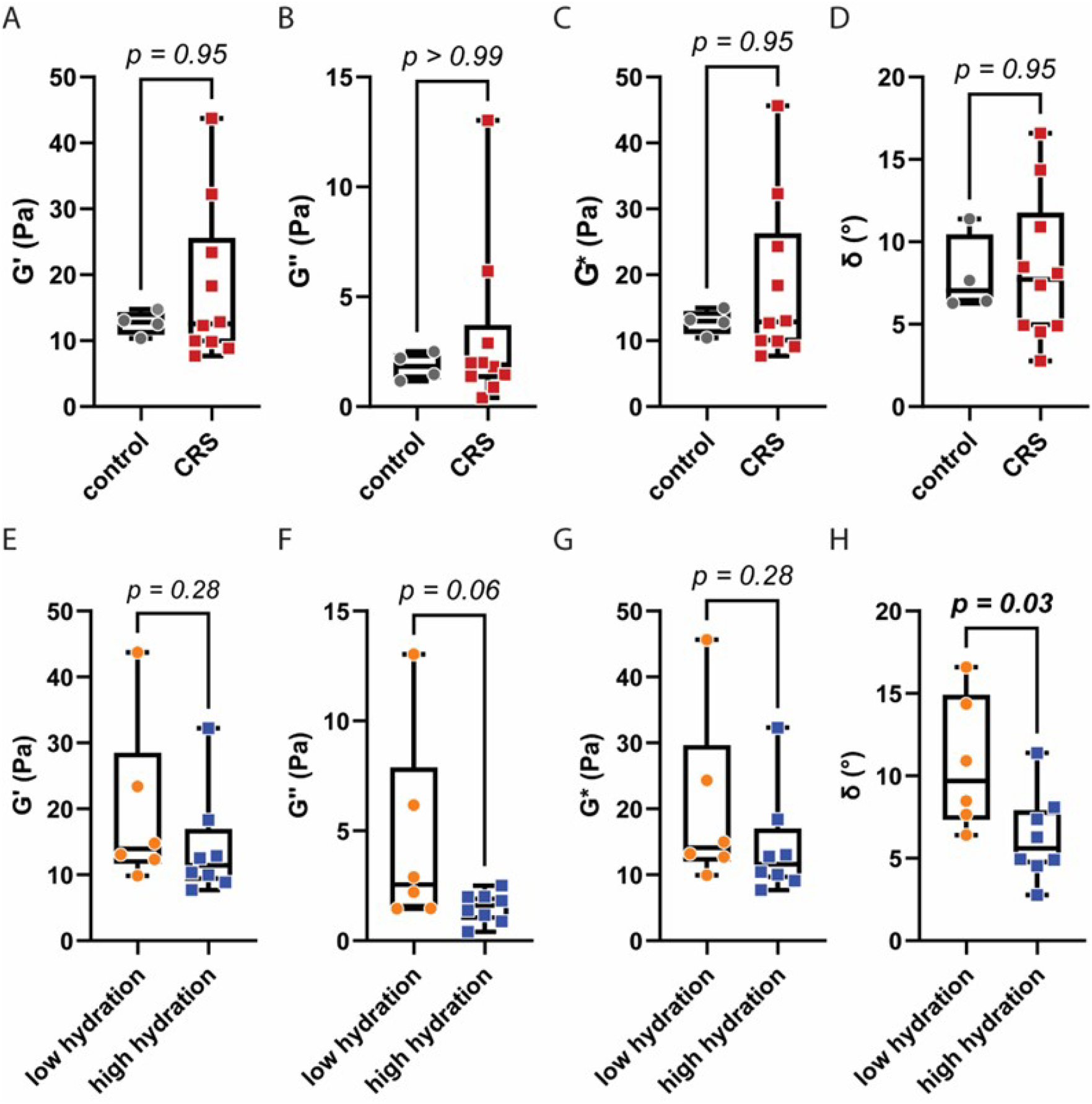
Mucus viscoelasticity by oscillatory shear rheology at 1 Hz. In A–D), CRS mucus (*n* = 10) and control mucus (*n* =4) are compared by A) elastic modulus; B) viscous modulus; C) complex modulus; and D) phase angle. In E–H), samples were stratified by high or low hydration (below vs. above cohort mean, *n* = 8 and *n* = 6, respectively) and compared by E) elastic modulus; F) viscous modulus; G) complex modulus; and H) phase angle. Each value is the average of four frequency sweeps. A significant decrease in H) phase angle was observed between low and high hydration (*p* =0.03). Mann-Whitney t-tests were performed to determine significance between groups.

**Figure 3.**
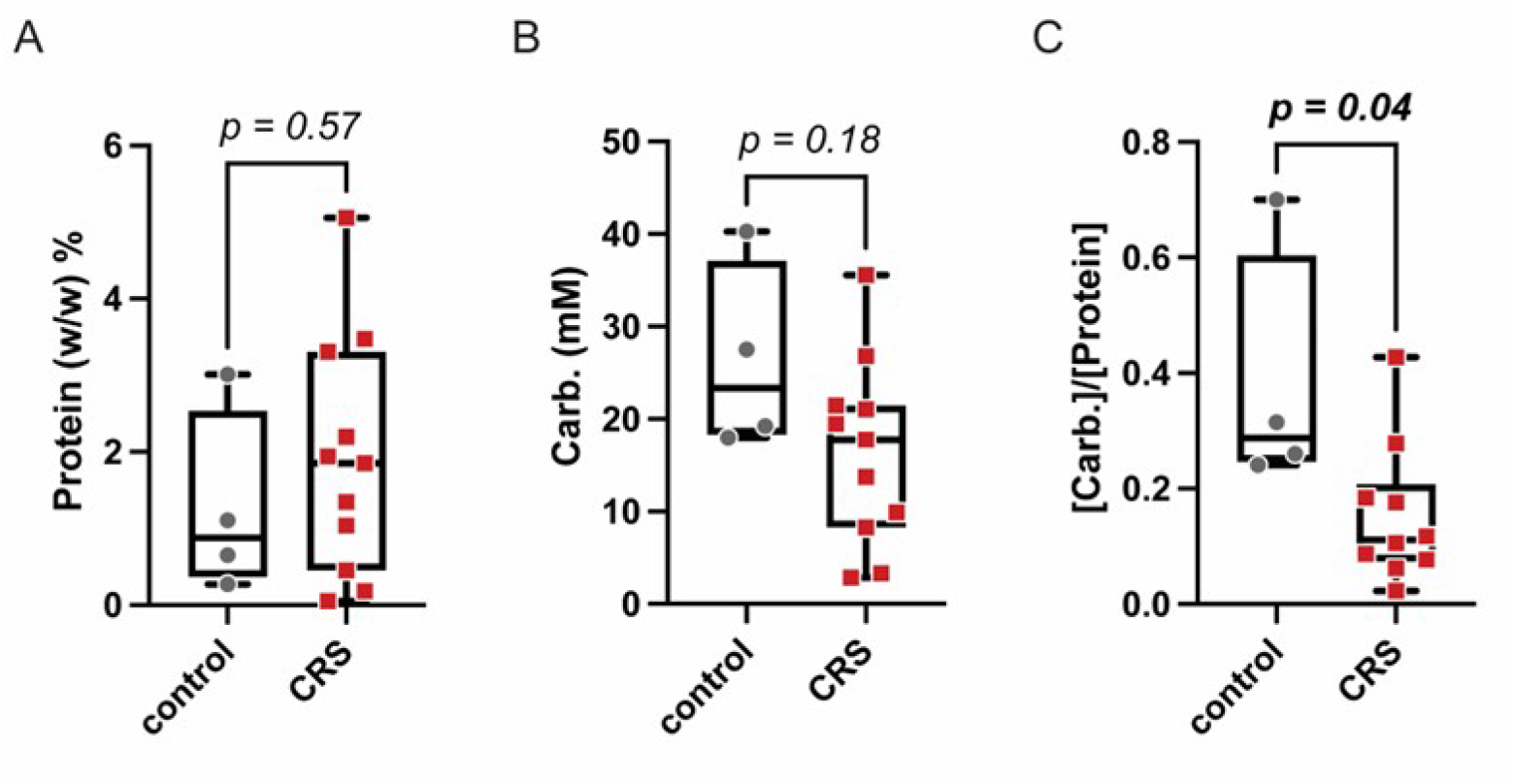
Protein and carbohydrate content of CRS mucus (*n* = 11) and control mucus (*n* =4) where A) is total protein (w/w %) obtained by BCA assay; B) is total carbohydrate content (mM) obtained by phenol-sulfuric acid assay; and C) is the mM ratio of carbohydrate to protein, which showed a significant decrease in CRS compared to controls (*p* = 0.04). Mann-Whitney t-tests were performed to determine significance between groups.

Stratification by hydration (below vs. above cohort mean) showed higher G′ in low hydration samples (19.5 ± 12.7, CV = 65.3% vs. 14.1 ± 8.0 Pa, CV = 56.9%; *p* = 0.28) as well as increased G″ (4.5 ± 4.5, CV = 99.4%; vs. 1.5 ± 0.7 Pa, CV = 45.2%; *p* = 0.06) (Figure 2E–H). G* followed a similar trend but was not significant (*p* = 0.28). In contrast, δ was significantly higher in low hydration samples (10.73 ± 4.02° vs. 6.29 ± 2.66°; *p* = 0.03).

### 3.4 Sinonasal mucus protein and carbohydrate content

Aside from water and ions, the primary component of mucus is mucin glycoproteins. Serine and threonine residues in the mucin backbone are covalently modified with carbohydrates ranging from a single monosaccharide to oligosaccharides of up to ~15 units.^2^ The genetic and metabolic processes regulating glycosylation remain poorly understood. Increasing saccharide density increases mucin peptide extension and rigidification as well as expansion of the hydration shell.^2,46^ These structural changes can influence overall mucus water-binding capacity and rheology, with downstream pathophysiological consequences.^45^

To assess protein and carbohydrate content, we quantified total protein and total carbohydrate content of hydrated mucus. For total protein, we used a BCA assay based on chelation of Cu^2+^ ions by the peptide backbone. Total carbohydrate content was determined using a phenol–sulfuric acid assay. Both assays are highly sensitive, well established, and provide straightforward colorimetric readouts. We note that full proteomics of CRS mucus has been reported elsewhere.^47^

Protein content of CRS mucus was higher than that of non-CRS mucus; however, this difference was not statistically significant (1.90 ± 1.55 w/w%, CV= 81.6% vs. 1.26 ± 1.22 w/w%, CV= 96.5%, respectively; *p* = 0.57). In contrast, the total carbohydrate content of CRS mucus (16.0 ± 0.001 mM) was lower than that of non-CRS mucus (25.9 ± 0.001 mM). This difference showed a stronger trend than for protein content but did not reach statistical significance (*p* = 0.18). Consistent with the hydration data, inter-sample variability in total carbohydrate content was greater in the CRS group than in the control group (CV = 62.65% vs. 39.67%, respectively). No comparisons reached statistical significance for stratification by polyp phenotype (see SI).

To further examine the relationship between protein and carbohydrate content, we calculated the molar ratio of carbohydrates to protein for each mucus sample. Protein w/w% was converted to molarity assuming an average amino acid molecular weight of 110 Da. Notably, the molar ratio of carbohydrates (monosaccharides) to protein (amino acids) was significantly lower in the CRS group (0.15 ± 0.12) compared to the control group (0.38 ± 0.22) (*p* = 0.04).

### 3.5 Analysis of mucus Sia and Fuc content

Carbohydrates influence mucin biophysics and biochemistry. Saccharides that terminate glycan chains, such as Sia and Fuc, are better poised for binding events than saccharides buried near the peptide backbone. These two sugars have well-characterized and diverse interactions with commensal and pathogenic microbes and immune cells.^34,35^ Further, anionic Sia can influence mucin polymer cross-linking through ionic bonds.^48^ We quantified mucus Sia and Fuc content in w/w% using well-established colorimetric assays. Fuc was measured via L-fucose dehydrogenase–mediated NADPH production^49,50^, and Sia via the Warren method following acid hydrolysis^51^. We also converted to Sia and Fuc to molar concentrations to calculate their ratios to total carbohydrate.

Fucose content did not differ between CRS and controls (1.17 ± 1.00 vs. 1.07 ± 0.86 w/w%; *p* = 0.95) (Fig. 4A), nor did the Fuc-to-carbohydrate ratio (0.45 ± 0.54 vs. 0.27 ± 0.17; *p* = 0.99) (Fig. 4B). Consistent with other measurements, CRS samples showed greater variability (CV = 119 % vs. 63.5%). No significant differences were observed when stratified by polyp status (Fig. 4E, F).

**Figure 4.**
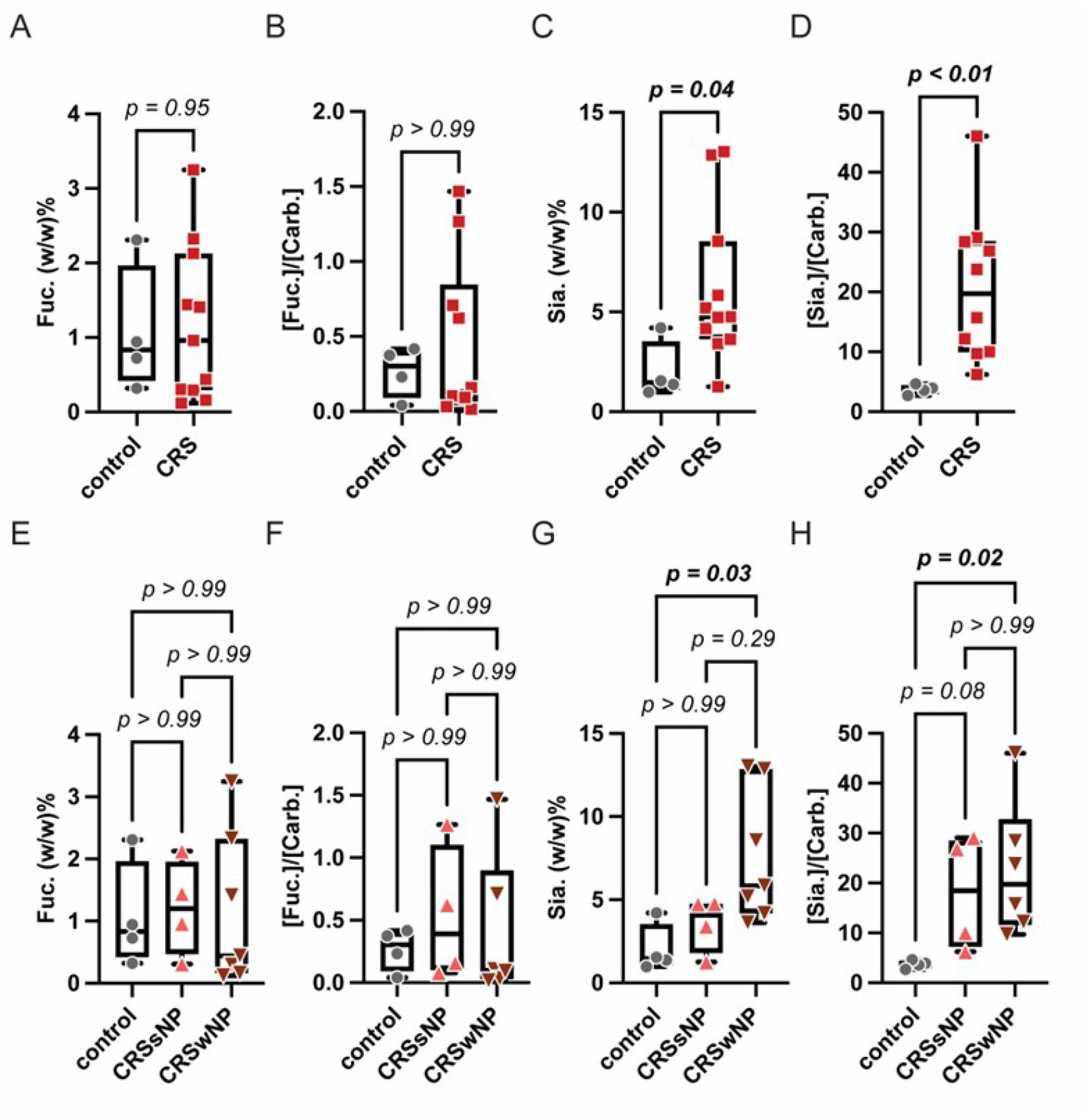
Glycan analysis of sialic acid and fucose content by chemical and enzymatic assays. A–D) are CRS mucus (*n* = 10) vs. control mucus (*n* =4) while E–F) compare CRSwNP (*n* = 7), CRSsNP (*n* = 4), and control mucus (*n* =4). A, E) are mucus fucose content as % of dehydrated mass; B, F) are the molar ratio of fucose to total carbohydrates; C, G) are mucus sialic acid content as % of dehydrated mass; and D, H) are the molar ratio of sialic acid to total carbohydrates. Mann-Whitney t-tests were performed to determine significance between groups.

In contrast, Sia content was significantly elevated in CRS mucus compared to controls (6.13 ± 3.81 vs. 2.03 ± 1.47 w/w%; *p* = 0.04) (Fig. 4C), with lower variability than Fuc (CV = 62.1% in CRS vs. 72.8% in controls). The Sia-to-carbohydrate ratio was also significantly elevated in CRS (20.8 ± 12.3 vs. 3.72 ± 0.80; *p* = 0.002) (Fig. 4D). Subgroup analysis indicated that elevated Sia levels were primarily driven by the CRSwNP subgroup (Fig. 4G, H). Sia in CRSwNP mucus was significantly higher than in the control by both w/w% (7.61 ± 3.97 vs. 2.03 ± 1.47 w/w%; *p* = 0.03) and by Sia-to-total carbohydrate ratio (22.6 ± 13.5 vs. 3.7 ± 0.80; *p* = 0.02). By w/w%, there was no statistically significant difference between the control samples and CRSsNP (*p* = 0.99). By Sia-to-total carbohydrate ratio, CRSsNP Sia levels were elevated as compared to control but only approached significance (18.1 ± 11.6; *p* = 0.08). Comparison of CRSsNP and CRSwNP by w/w% or Sia-to-total carbohydrate ratio did not reveal significant differences (*p* = 0.29; *p* = 0.99, respectively).

## Discussion

This study provides a quantitative compositional analysis of CRS mucus. Prior work has relied largely on tissue-based assessments or qualitative glycan profiling, whereas we directly quantified mucus-phase properties, including hydration, rheology, and relative abundances of cations, protein, carbohydrates, and specific glycans. Across methods, CRS mucus generally exhibited greater heterogeneity than controls. Although limited by sample size, these findings support coordinated alterations in ionic environment and mucin glycosylation in CRS.

Cation content was examined by ultra-sensitive ICP-QQQ and was elevated in CRS for both monovalent and divalent species, with approximately twofold higher Ca^2+^ and Mg^2+^. While prior direct ion quantitation in CRS mucus is limited, the overall hierarchy aligns with healthy nasal secretions.^52^ A murine CRS model indicated similarly altered K^+^ and Na^+^, though methodological and species differences constrain comparison.^53^ Broadly, sinonasal epithelial ion transport studies implicate disrupted ionic flux in impaired MCC and mucosal dysfunction.^26,54^ Our data extend this framework by suggesting that mucus itself may function as an ion reservoir. Further, small changes in ion-mucin interactions such as electrostatic screening by monovalent cations and crosslinking by divalent ions, may disproportionately contribute to altered mucosal rheology, transport, and stasis. ^2,9,55,56^

Rheologic measurements confirmed characteristic viscoelastic signatures in both CRS and control groups. Lower mucus water content increased viscoelasticity, most evident in phase angle with minor changes in viscous moduli. CRS samples showed greater variability and trended toward increased stiffness and resistance, consistent with other airway diseases.^45,57,58^ This heterogeneity likely reflects differences in mucin concentration, structure, and intermolecular interactions beyond hydration and ionic effects. One limitation is that macro-rheology captures bulk properties but may miss microscale variation in mesh size and network structure that can only be captured using micro-rheology techniques.

The significantly reduced carbohydrate-to-protein ratio in CRS mucus indicates altered composition and suggests mucin hypoglycosylation, which has been reported in other inflammatory diseases and cancers.^59–61^ Although other macromolecules may contribute to this ratio, mucins dominate mucus dry mass.^2,10^ Advanced mass spectrometry techniques should be applied in future work to fully characterize the glycans. However, we refined the picture through quantification of mucin terminal glycans, Sia and Fuc, that influence mucus biophysics and biochemistry. Fuc did not differ significantly in CRS, consistent with a prior report.^36^ High inter-sample variability observed in our cohort likely reflects microbial influences.^62^

In contrast, elevated sialylation distinguished CRS mucus. This aligns with several studies ^33–36^, though there are discrepancies in tissue vs. mucus sampling and limitations of lectin-based methods known to have poor specificity and high glycan cross-reactivity^37,63–65^. The quantitative assays used here improve specificity and sensitivity, but we note the techniques do not resolve linkage types. Subgroup analysis indicated that glycosylation changes were most pronounced in CRSwNP, consistent with its distinct inflammatory and epithelial profiles.^66^ Increased sialylation in this subgroup may reflect differences in metabolic, microbial, or immune activity, and should be examined further.

Increased sialylation and reduced carbohydrate-to-protein ratio together reinforce likely mucin hypoglycosylation since Sia typically terminates oligosaccharide chain growth. Increased sialylation also raises mucin charge density and promotes polymer expansion and hydration.^2,67^ However, elevated cations can screen these interactions, collapse networks, and increase viscoelasticity.^54,68,69^ The observed concurrent changes in cation content and sialylation are consistent with this interplay and provide a mechanistic basis for alterations in MCC in CRS.

Hypersialylation may also influence microbial and immune interactions relevant to CRS. Sia is a direct ligand for diverse pathogens^67^ and indeed elevated Sia has been linked to biofilm burden in CRS^34,35^. Further, sialic acid–binding immunoglobulin-like lectin receptors (Siglecs) are broadly expressed on leukocytes and binding typically downregulates their activity.^70^ Changes in mucin glycosylation could therefore shape host–microbe and immune dynamics within the sinonasal environment, highlighting a potential therapeutic target.^71–73^

This study is limited by its relatively small sample size, particularly with respect to CRSsNP and CRSwNP subgroups, which reduces statistical power and could limit detection of more nuanced mucus compositional differences. Since the study was conducted at a single institution in a cross-sectional manner, the generalizability of the findings to broader CRS populations or to temporal fluctuations may be limited. Although our methodology improved specificity for detection of sialic acid and fucose, this is only a partial view of mucin glycosylation and full glycomics profiling should be undertaken in future work.

Clinically, our collective findings suggest refocusing to include consideration of mucus structure and composition alongside mucus quantity. Our data indicate that CRS mucus reflects coupled disruptions in ionic regulation and mucin glycosylation consistent with other airway disorders.^2,3,9^ Targeting these biophysical and biochemical abnormalities may be required to restore effective MCC, reduce inflammation, and meaningfully modify disease course.

## Conclusion

In this study we characterized CRS mucus including hydration, ionic content, rheology, protein, and glycan composition and stratified data by CRSsNP and CRSwCP. We identified increased cation content, increased sialylation, and decreased carbohydrate-to-protein ratios as characteristics of CRS. These altered ratios, in the absence of large changes in bulk protein or total carbohydrate, support a model of selective glycan and ion remodeling beyond generalized hypersecretion. By linking these compositional changes to established principles of mucin gel behavior, we provide a mechanistic bridge between molecular alterations and clinically relevant mucus dysfunction. This study advances current understanding of CRS pathophysiology, highlighting mucin glycosylation as a potential contributor to impaired mucus clearance and as a target for future therapeutics.

## Data Availability

All data produced in the present work are contained in the manuscript

