## Supplemental Material for "Cation Enrichment and Hypersialylation in Chronic Rhinosinusitis Mucus"

ł indicates equal contribution

**Table of Contents**

**1. Supplemental Figures**

Figure S1. Mucus hydration by polyp phenotype

Figure S2. Mucus cation content by polyp phenotype

Figure S3. Mucus viscoelasticity at 1 Hz by polyp phenotype

Figure S4. Mucus viscoelasticity at 10 Hz, combined CRS and by polyp phenotype

Figure S5. Mucus viscoelasticity at 10 Hz by hydration status

Figure S6. Mucus total protein and carbohydrate content by polyp phenotype

**2. Data Analysis and Statistical Tables**

Table 1. Summary of p-values

Table 2. Summary of descriptive statistics

**3. Detailed Rheological Procedures**

**4. Detailed Assay Procedures**

4.1 Total protein assay detailed procedure

4.2 Total carbohydrate assay detailed procedure

4.3 Fucose assay detailed procedure

4.4 Sialic acid assay detailed procedure

### 1. Supplemental Figures

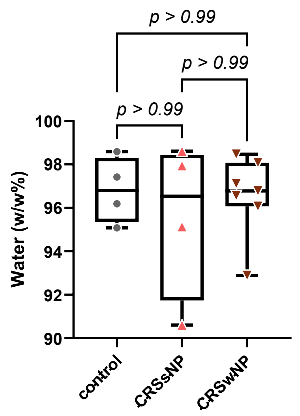

**Figure S1**: Hydration (w/w %) of CRSwNP mucus (*n* = 7), CRSsNP mucus (*n* = 4), and control mucus (*n* =4) by lyophilization mass loss. Mann-Whitney t-tests were performed to determine significance between groups.

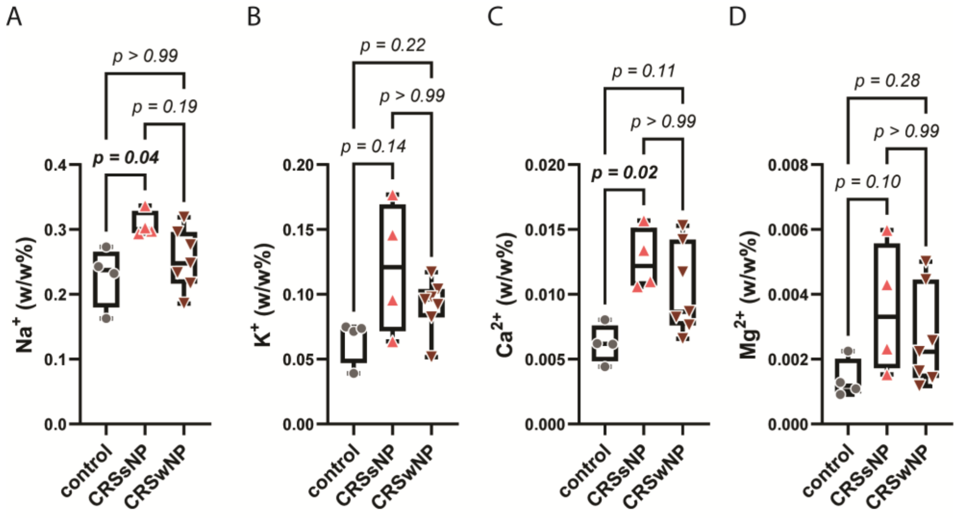

**Figure S2**: Cation content (w/w %) of CRSwNP mucus ($n$ = 7), CRSsNP mucus ($n$ = 4), and control mucus ($n$ =4) by ICP-QQQ. Mann-Whitney t-tests were performed to determine significance between groups.

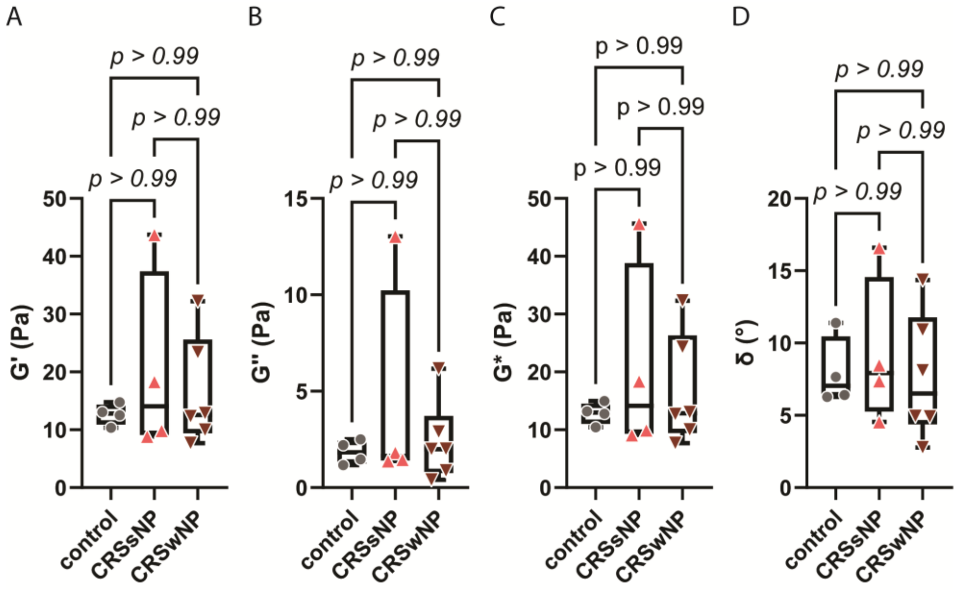

**Figure S3**: Mucus viscoelasticity by oscillatory shear rheology at 1 Hz. In A–D), CRSwNP mucus ($n$ = 7), CRSsNP mucus ($n$ = 4), and control mucus ($n$ =4) are compared by A) elastic modulus; B) viscous modulus; C) complex modulus; and D) phase angle. Mann-Whitney t-tests were performed to determine significance between groups.

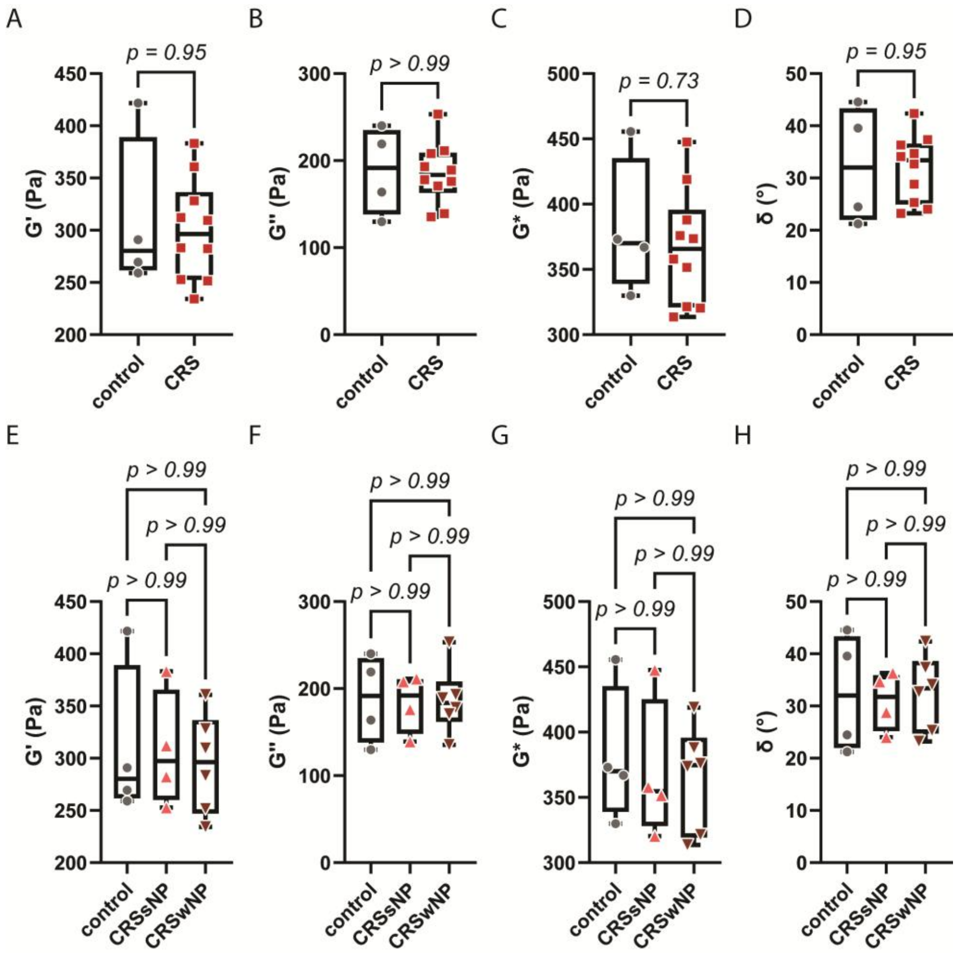

**Figure S4**: Mucus viscoelasticity by oscillatory shear rheology at 10 Hz. In A–D), CRS mucus (*n* = 10) and control mucus (*n* =4) are compared, while in E–H), CRSwNP mucus (*n* = 7), CRSsNP mucus (*n* = 4), and control mucus (*n* =4) are compared. A, E) elastic modulus; B, F) viscous modulus; C, G) complex modulus; and D, H) phase angle. Mann-Whitney t-tests were performed to determine significance between groups.

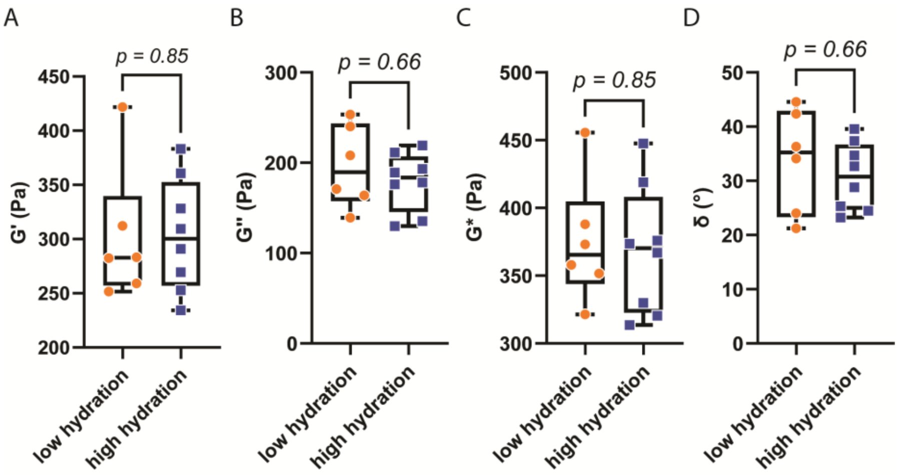

**Figure S5**: Mucus viscoelasticity by oscillatory shear rheology at 10 Hz. Samples were stratified by high or low hydration (below vs. above cohort mean, $n$ = 8 and $n$ = 6, respectively) and compared by A) elastic modulus; B) viscous modulus; C) complex modulus; and D) phase angle. Each value is the average of four frequency sweeps. Mann-Whitney t-tests were performed to determine significance between groups.

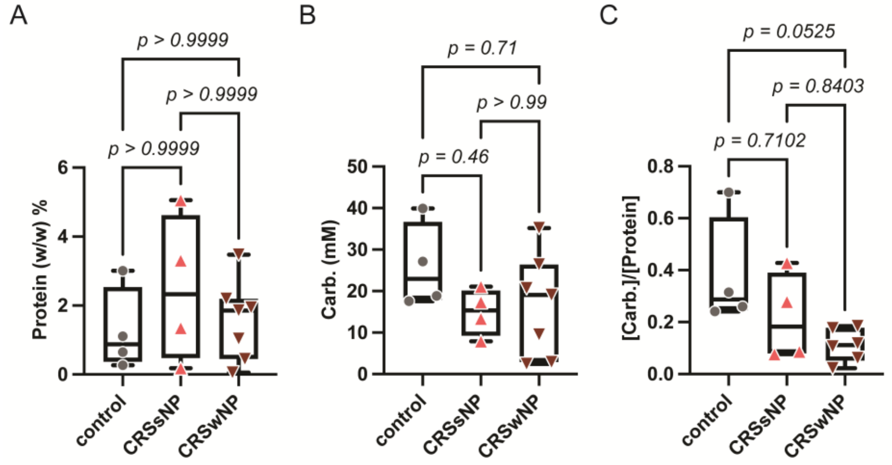

**Figure S6**: Protein and carbohydrate content of CRSwNP mucus ($n$ = 7), CRSsNP mucus ($n$ = 4), and control mucus ($n$ =4) where A) is total protein (w/w %) obtained by BCA assay; B) is total carbohydrate content (mM) obtained by phenol-sulfuric acid assay; and C) is the mM ratio of carbohydrate to protein. Mann-Whitney t-tests were performed to determine significance between groups.

### 2. Data Analysis and Statistical Tables

Commented [JK1]: Update tables

**Table 1.** Summary of p-values for differences between groups by Kruskal-Wallis, after Dunn's multiple comparisons test (MCT), or Mann-Whitney. Statistical significance is denoted * for p < 0.05.

| Hydration (w/w%) | | |
|---|---|---|
| Comparison | p-value | test |
| CRSsNP vs. CRSwNP vs. control | 0.9869 | Kruskal-Wallis |
| CRSsNP vs. CRSwNP | >0.9999 | Dunn's MCT from Kruskal-Wallis |
| CRSsNP vs. control | >0.9999 | Dunn's MCT from Kruskal-Wallis |
| CRSwNP vs. control | >0.9999 | Dunn's MCT from Kruskal-Wallis |
| CRS vs. control | 0.9495 | Mann-Whitney |
| CRSsNP vs. CRSwNP | 0.9273 | Mann-Whitney |

| Total carbohydrate content (mM) | | |
|---|---|---|
| Comparison | p-value | test |
| CRSsNP vs. CRSwNP vs. control | 0.3505 | Kruskal-Wallis |
| CRSsNP vs. CRSwNP | >0.9999 | Dunn's MCT from Kruskal-Wallis |
| CRSsNP vs. control | 0.4642 | Dunn's MCT from Kruskal-Wallis |
| CRSwNP vs. control | 0.7081 | Dunn's MCT from Kruskal-Wallis |
| CRS vs. control | 0.1773 | Mann-Whitney |
| CRSsNP vs. CRSwNP | 0.9273 | Mann-Whitney |

| Total protein content (w/w%) by dry mass | | |
|---|---|---|
| Comparison | p-value | test |
| CRSsNP vs. CRSwNP vs. control | 0.5157 | Kruskal-Wallis |
| CRSsNP vs. CRSwNP | >0.9999 | Dunn's MCT from Kruskal-Wallis |
| CRSsNP vs. control | 0.707 | Dunn's MCT from Kruskal-Wallis |
| CRSwNP vs. control | >0.9999 | Dunn's MCT from Kruskal-Wallis |
| CRS vs. control | 0.4117 | Mann-Whitney |
| CRSsNP vs. CRSwNP | 0.4121 | Mann-Whitney |

| Total protein content (w/w%) by hydrated mass | | |
|---|---|---|
| Comparison | p-value | test |
| CRSsNP vs. CRSwNP vs. control | 0.7091 | Kruskal-Wallis |
| CRSsNP vs. CRSwNP | >0.9999 | Dunn's MCT from Kruskal-Wallis |
| CRSsNP vs. control | >0.9999 | Dunn's MCT from Kruskal-Wallis |
| CRSwNP vs. control | >0.9999 | Dunn's MCT from Kruskal-Wallis |
| CRS vs. control | 0.5714 | Mann-Whitney |
| CRSsNP vs. CRSwNP | 0.6485 | Mann-Whitney |

| Carbohydrate to protein ratio (AU) | | |
|---|---|---|
| Comparison | p-value | test |
| CRSsNP vs. CRSwNP vs. control | 0.0505 | Kruskal-Wallis |
| CRSsNP vs. CRSwNP | 0.7102 | Dunn's MCT from Kruskal-Wallis |
| CRSsNP vs. control | 0.0525 | Dunn's MCT from Kruskal-Wallis |
| CRSwNP vs. control | 0.8403 | Dunn's MCT from Kruskal-Wallis |
| CRS vs. control | 0.036* | Mann-Whitney |
| CRSsNP vs. CRSwNP | 0.4762 | Mann-Whitney |

| Fucose (w/w%) by dry mass | | |
|---|---|---|
| Comparison | p-value | test |

| | | |
|---|---|---|
| CRSsNP vs. CRSwNP vs. control | 0.8463 | Kruskal-Wallis |
| CRSsNP vs. CRSwNP | >0.9999 | Dunn's MCT from Kruskal-Wallis |
| CRSsNP vs. control | >0.9999 | Dunn's MCT from Kruskal-Wallis |
| CRSwNP vs. control | >0.9999 | Dunn's MCT from Kruskal-Wallis |
| CRS vs. control | 0.9495 | Mann-Whitney |
| CRSsNP vs. CRSwNP | 0.6485 | Mann-Whitney |

Fucose (w/w%) by hydrated mass

| Comparison | p-value | test |
|---|---|---|
| CRSsNP vs. CRSwNP vs. control | 0.8463 | Kruskal-Wallis |
| CRSsNP vs. CRSwNP | >0.9999 | Dunn's MCT from Kruskal-Wallis |
| CRSsNP vs. control | >0.9999 | Dunn's MCT from Kruskal-Wallis |
| CRSwNP vs. control | >0.9999 | Dunn's MCT from Kruskal-Wallis |
| CRS vs. control | 0.9495 | Mann-Whitney |
| CRSsNP vs. CRSwNP | 0.6485 | Mann-Whitney |

Fucose (mM)

| Comparison | p-value | test |
|---|---|---|
| CRSsNP vs. CRSwNP vs. control | 0.5668 | Kruskal-Wallis |
| CRSsNP vs. CRSwNP | >0.9999 | Dunn's MCT from Kruskal-Wallis |
| CRSsNP vs. control | >0.9999 | Dunn's MCT from Kruskal-Wallis |
| CRSwNP vs. control | 0.8364 | Dunn's MCT from Kruskal-Wallis |
| CRS vs. control | 0.4117 | Mann-Whitney |
| CRSsNP vs. CRSwNP | 0.6485 | Mann-Whitney |

% of total carbohydrates that are fucose

| Comparison | p-value | test |
|---|---|---|
| CRSsNP vs. CRSwNP vs. control | 0.9312 | Kruskal-Wallis |
| CRSsNP vs. CRSwNP | >0.9999 | Dunn's MCT from Kruskal-Wallis |
| CRSsNP vs. control | >0.9999 | Dunn's MCT from Kruskal-Wallis |
| CRSwNP vs. control | >0.9999 | Dunn's MCT from Kruskal-Wallis |
| CRS vs. control | 0.8513 | Mann-Whitney |
| CRSsNP vs. CRSwNP | 0.9273 | Mann-Whitney |

Sialic acid (w/w%) by dry mass

| Comparison | p-value | test |
|---|---|---|
| CRSsNP vs. CRSwNP vs. control | 0.0176* | Kruskal-Wallis |
| CRSsNP vs. CRSwNP | 0.293 | Dunn's MCT from Kruskal-Wallis |
| CRSsNP vs. control | >0.9999 | Dunn's MCT from Kruskal-Wallis |
| CRSwNP vs. control | 0.0325* | Dunn's MCT from Kruskal-Wallis |
| CRS vs. control | 0.0396* | Mann-Whitney |
| CRSsNP vs. CRSwNP | 0.0727 | Mann-Whitney |

Sialic acid (w/w%) by hydrated mass

| Comparison | p-value | test |
|---|---|---|
| CRSsNP vs. CRSwNP vs. control | 0.1611 | Kruskal-Wallis |
| CRSsNP vs. CRSwNP | >0.9999 | Dunn's MCT from Kruskal-Wallis |
| CRSsNP vs. control | 0.707 | Dunn's MCT from Kruskal-Wallis |
| CRSwNP vs. control | 0.1679 | Dunn's MCT from Kruskal-Wallis |
| CRS vs. control | 0.0777 | Mann-Whitney |
| CRSsNP vs. CRSwNP | 0.7879 | Mann-Whitney |

Sialic acid (mM)

| Comparison | p-value | test |
|---|---|---|
| CRSsNP vs. CRSwNP vs. control | 0.038* | Kruskal-Wallis |
| CRSsNP vs. CRSwNP | >0.9999 | Dunn's MCT from Kruskal-Wallis |
| CRSsNP vs. control | 0.3992 | Dunn's MCT from Kruskal-Wallis |
| CRSwNP vs. control | 0.0403* | Dunn's MCT from Kruskal-Wallis |
| CRS vs. control | 0.0176* | Mann-Whitney |
| CRSsNP vs. CRSwNP | 0.5273 | Mann-Whitney |

| % of total carbohydrates that are sialic acid | | |
|---|---|---|
| Comparison | p-value | test |
| CRSsNP vs. CRSwNP vs. control | 0.0074* | Kruskal-Wallis |
| CRSsNP vs. CRSwNP | >0.9999 | Dunn's MCT from Kruskal-Wallis |
| CRSsNP vs. control | 0.084 | Dunn's MCT from Kruskal-Wallis |
| CRSwNP vs. control | 0.0198* | Dunn's MCT from Kruskal-Wallis |
| CRS vs. control | 0.002* | Mann-Whitney |
| CRSsNP vs. CRSwNP | 0.7619 | Mann-Whitney |

| Sodium (w/w%) by hydrated mass | | |
|---|---|---|
| Comparison | p-value | test |
| CRSsNP vs. CRSwNP vs. control | 0.032* | Kruskal-Wallis |
| CRSsNP vs. CRSwNP | 0.1886 | Dunn's MCT from Kruskal-Wallis |
| CRSsNP vs. control | 0.0428* | Dunn's MCT from Kruskal-Wallis |
| CRSwNP vs. control | >0.9999 | Dunn's MCT from Kruskal-Wallis |
| CRS vs. control | 0.104 | Mann-Whitney |
| CRSsNP vs. CRSwNP | 0.0727 | Mann-Whitney |

| Magnesium (w/w%) by hydrated mass | | |
|---|---|---|
| Comparison | p-value | test |
| CRSsNP vs. CRSwNP vs. control | 0.0854 | Kruskal-Wallis |
| CRSsNP vs. CRSwNP | >0.9999 | Dunn's MCT from Kruskal-Wallis |
| CRSsNP vs. control | 0.0984 | Dunn's MCT from Kruskal-Wallis |
| CRSwNP vs. control | 0.2778 | Dunn's MCT from Kruskal-Wallis |
| CRS vs. control | 0.0396* | Mann-Whitney |
| CRSsNP vs. CRSwNP | 0.5273 | Mann-Whitney |

| Potassium (w/w%) by hydrated mass | | |
|---|---|---|
| Comparison | p-value | test |
| CRSsNP vs. CRSwNP vs. control | 0.0994 | Kruskal-Wallis |
| CRSsNP vs. CRSwNP | >0.9999 | Dunn's MCT from Kruskal-Wallis |
| CRSsNP vs. control | 0.1443 | Dunn's MCT from Kruskal-Wallis |
| CRSwNP vs. control | 0.2234 | Dunn's MCT from Kruskal-Wallis |
| CRS vs. control | 0.0396* | Mann-Whitney |
| CRSsNP vs. CRSwNP | 0.5273 | Mann-Whitney |

| Calcium (w/w%) by hydrated mass | | |
|---|---|---|
| Comparison | p-value | test |
| CRSsNP vs. CRSwNP vs. control | 0.0114* | Kruskal-Wallis |
| CRSsNP vs. CRSwNP | >0.9999 | Dunn's MCT from Kruskal-Wallis |
| CRSsNP vs. control | 0.0216* | Dunn's MCT from Kruskal-Wallis |
| CRSwNP vs. control | 0.11 | Dunn's MCT from Kruskal-Wallis |
| CRS vs. control | 0.0059* | Mann-Whitney |
| CRSsNP vs. CRSwNP | 0.3152 | Mann-Whitney |

| Sodium (w/w%) by dry mass | | |
|---|---|---|
| Comparison | p-value | test |
| CRSsNP vs. CRSwNP vs. control | 0.9767 | Kruskal-Wallis |
| CRSsNP vs. CRSwNP | >0.9999 | Dunn's MCT from Kruskal-Wallis |
| CRSsNP vs. control | >0.9999 | Dunn's MCT from Kruskal-Wallis |
| CRSwNP vs. control | >0.9999 | Dunn's MCT from Kruskal-Wallis |
| CRS vs. control | 0.8513 | Mann-Whitney |
| CRSsNP vs. CRSwNP | >0.9999 | Mann-Whitney |

| Magnesium (w/w%) by dry mass | | |
|---|---|---|
| Comparison | p-value | test |
| CRSsNP vs. CRSwNP vs. control | 0.0527 | Kruskal-Wallis |
| CRSsNP vs. CRSwNP | >0.9999 | Dunn's MCT from Kruskal-Wallis |
| CRSsNP vs. control | 0.0656 | Dunn's MCT from Kruskal-Wallis |
| CRSwNP vs. control | 0.2297 | Dunn's MCT from Kruskal-Wallis |
| CRS vs. control | 0.0264* | Mann-Whitney |
| CRSsNP vs. CRSwNP | 0.4121 | Mann-Whitney |

| Potassium (w/w%) by dry mass | | |
|---|---|---|
| Comparison | p-value | test |
| CRSsNP vs. CRSwNP vs. control | 0.8834 | Kruskal-Wallis |
| CRSsNP vs. CRSwNP | >0.9999 | Dunn's MCT from Kruskal-Wallis |
| CRSsNP vs. control | >0.9999 | Dunn's MCT from Kruskal-Wallis |
| CRSwNP vs. control | >0.9999 | Dunn's MCT from Kruskal-Wallis |
| CRS vs. control | 0.6608 | Mann-Whitney |
| CRSsNP vs. CRSwNP | >0.9999 | Mann-Whitney |

| Calcium (w/w%) by dry mass | | |
|---|---|---|
| Comparison | p-value | test |
| CRSsNP vs. CRSwNP vs. control | 0.5764 | Kruskal-Wallis |
| CRSsNP vs. CRSwNP | >0.9999 | Dunn's MCT from Kruskal-Wallis |
| CRSsNP vs. control | 0.9122 | Dunn's MCT from Kruskal-Wallis |
| CRSwNP vs. control | >0.9999 | Dunn's MCT from Kruskal-Wallis |
| CRS vs. control | 0.3429 | Mann-Whitney |
| CRSsNP vs. CRSwNP | 0.9273 | Mann-Whitney |

| Elastic Modulus, G’ @ 1 Hz (Pa) | | |
|---|---|---|
| Comparison | p-value | test |
| CRSsNP vs. CRSwNP vs. control | 0.9966 | Kruskal-Wallis |
| CRSsNP vs. CRSwNP | >0.9999 | Dunn's MCT from Kruskal-Wallis |
| CRSsNP vs. control | >0.9999 | Dunn's MCT from Kruskal-Wallis |
| CRSwNP vs. control | >0.9999 | Dunn's MCT from Kruskal-Wallis |
| CRS vs. control | 0.9451 | Mann-Whitney |
| CRSsNP vs. CRSwNP | >0.9999 | Mann-Whitney |
| Low vs. high hydration | 0.2824 | Mann-Whitney |

| Viscous Modulus, G” @ 1 Hz (Pa) | | |
|---|---|---|
| Comparison | p-value | test |
| CRSsNP vs. CRSwNP vs. control | >0.9999 | Kruskal-Wallis |
| CRSsNP vs. CRSwNP | >0.9999 | Dunn's MCT from Kruskal-Wallis |
| CRSsNP vs. control | >0.9999 | Dunn's MCT from Kruskal-Wallis |
| CRSwNP vs. control | >0.9999 | Dunn's MCT from Kruskal-Wallis |
| CRS vs. control | >0.9999 | Mann-Whitney |

| | | |
|---|---|---|
| CRSsNP vs. CRSwNP | >0.9999 | Mann-Whitney |
| Low vs. high hydration | 0.0593 | Mann-Whitney |

| Complex Modulus, G* @ 1 Hz (Pa) | | |
|---|---|---|
| Comparison | p-value | test |
| CRSsNP vs. CRSwNP vs. control | 0.9966 | Kruskal-Wallis |
| CRSsNP vs. CRSwNP | >0.9999 | Dunn's MCT from Kruskal-Wallis |
| CRSsNP vs. control | >0.9999 | Dunn's MCT from Kruskal-Wallis |
| CRSwNP vs. control | >0.9999 | Dunn's MCT from Kruskal-Wallis |
| CRS vs. control | 0.9451 | Mann-Whitney |
| CRSsNP vs. CRSwNP | >0.9999 | Mann-Whitney |
| Low vs. high hydration | 0.2824 | Mann-Whitney |

| Phase Angle, δ @ 1 Hz (°) | | |
|---|---|---|
| Comparison | p-value | test |
| CRSsNP vs. CRSwNP vs. control | 0.8849 | Kruskal-Wallis |
| CRSsNP vs. CRSwNP | >0.9999 | Dunn's MCT from Kruskal-Wallis |
| CRSsNP vs. control | >0.9999 | Dunn's MCT from Kruskal-Wallis |
| CRSwNP vs. control | >0.9999 | Dunn's MCT from Kruskal-Wallis |
| CRS vs. control | 0.9451 | Mann-Whitney |
| CRSsNP vs. CRSwNP | 0.7619 | Mann-Whitney |
| Low vs. high hydration | 0.0293* | Mann-Whitney |

| Elastic Modulus, G’ @ 10 Hz (Pa) | | |
|---|---|---|
| Comparison | p-value | test |
| CRSsNP vs. CRSwNP vs. control | 0.9417 | Kruskal-Wallis |
| CRSsNP vs. CRSwNP | >0.9999 | Dunn's MCT from Kruskal-Wallis |
| CRSsNP vs. control | >0.9999 | Dunn's MCT from Kruskal-Wallis |
| CRSwNP vs. control | >0.9999 | Dunn's MCT from Kruskal-Wallis |
| CRS vs. control | 0.9451 | Mann-Whitney |
| CRSsNP vs. CRSwNP | 0.7619 | Mann-Whitney |
| Low vs. high hydration | 0.8518 | Mann-Whitney |

| Viscous Modulus, G” @ 10 Hz (Pa) | | |
|---|---|---|
| Comparison | p-value | test |
| CRSsNP vs. CRSwNP vs. control | >0.9999 | Kruskal-Wallis |
| CRSsNP vs. CRSwNP | >0.9999 | Dunn's MCT from Kruskal-Wallis |
| CRSsNP vs. control | >0.9999 | Dunn's MCT from Kruskal-Wallis |
| CRSwNP vs. control | >0.9999 | Dunn's MCT from Kruskal-Wallis |
| CRS vs. control | >0.9999 | Mann-Whitney |
| CRSsNP vs. CRSwNP | 0.9143 | Mann-Whitney |
| Low vs. high hydration | 0.662 | Mann-Whitney |

| Complex Modulus, G* @ 10 Hz (Pa) | | |
|---|---|---|
| Comparison | p-value | test |
| CRSsNP vs. CRSwNP vs. control | 0.852 | Kruskal-Wallis |
| CRSsNP vs. CRSwNP | >0.9999 | Dunn's MCT from Kruskal-Wallis |
| CRSsNP vs. control | >0.9999 | Dunn's MCT from Kruskal-Wallis |
| CRSwNP vs. control | >0.9999 | Dunn's MCT from Kruskal-Wallis |
| CRS vs. control | 0.7333 | Mann-Whitney |
| CRSsNP vs. CRSwNP | 0.9143 | Mann-Whitney |
| Low vs. high hydration | 0.8518 | Mann-Whitney |

| Phase Angle, δ @ 10 Hz (°) | | |
|---|---|---|
| Comparison | p-value | test |
| CRSsNP vs. CRSwNP vs. control | 0.9722 | Kruskal-Wallis |
| CRSsNP vs. CRSwNP | >0.9999 | Dunn's MCT from Kruskal-Wallis |
| CRSsNP vs. control | >0.9999 | Dunn's MCT from Kruskal-Wallis |
| CRSwNP vs. control | >0.9999 | Dunn's MCT from Kruskal-Wallis |
| CRS vs. control | 0.9451 | Mann-Whitney |
| CRSsNP vs. CRSwNP | 0.9143 | Mann-Whitney |
| Low vs. high hydration | 0.662 | Mann-Whitney |

**Table 2.** Summary of descriptive statistics.

| Hydration (w/w%) | | | | |
|---|---|---|---|---|
| | control | CRS | CRSsNP | CRSwNP |
| Number of samples (n) | 4 | 11 | 4 | 7 |
| Minimum | 95.08 | 90.6 | 90.6 | 92.89 |
| Maximum | 98.59 | 98.61 | 98.61 | 98.47 |
| Range | 3.512 | 8.011 | 8.011 | 5.582 |
| Mean | 96.82 | 96.21 | 95.57 | 96.57 |
| Std. Deviation | 1.519 | 2.496 | 3.64 | 1.827 |
| Std. Error of Mean | 0.7596 | 0.7525 | 1.82 | 0.6905 |
| Lower 95% CI of mean | 94.4 | 94.53 | 89.78 | 94.88 |
| Upper 95% CI of mean | 99.23 | 97.88 | 101.4 | 98.26 |
| Coefficient of Variation | 1.569% | 2.594% | 3.809% | 1.892% |

| Total carbohydrate content (mM) | | | | |
|---|---|---|---|---|
| | control | CRS | CRSsNP | CRSwNP |
| Number of samples (n) | 4 | 11 | 4 | 7 |
| Minimum | 17.61 | 2.474 | 7.901 | 2.474 |
| Maximum | 39.91 | 35.21 | 21.1 | 35.21 |
| Range | 22.3 | 32.73 | 13.19 | 32.73 |
| Mean | 25.88 | 16.01 | 14.93 | 16.62 |
| Std. Deviation | 10.27 | 10.03 | 5.654 | 12.26 |
| Std. Error of Mean | 5.133 | 3.023 | 2.827 | 4.635 |
| Lower 95% CI of mean | 9.544 | 9.269 | 5.933 | 5.279 |
| Upper 95% CI of mean | 42.21 | 22.74 | 23.93 | 27.96 |
| Coefficient of Variation | 39.67% | 62.65% | 37.87% | 73.78% |

| Total protein content (w/w%) by hydrated mass | | | | |
|---|---|---|---|---|
| | control | CRS | CRSsNP | CRSwNP |
| Number of samples (n) | 4 | 11 | 4 | 7 |
| Minimum | 0.2719 | 0.05408 | 0.1828 | 0.05408 |
| Maximum | 3.013 | 5.06 | 5.06 | 3.477 |
| Range | 2.741 | 5.005 | 4.877 | 3.423 |
| Mean | 1.261 | 1.901 | 2.475 | 1.573 |

| | | | | |
|---|---|---|---|---|
| Std. Deviation | 1.217 | 1.551 | 2.152 | 1.161 |
| Std. Error of Mean | 0.6084 | 0.4675 | 1.076 | 0.4387 |
| Lower 95% CI of mean | -0.6752 | 0.8595 | -0.9493 | 0.4999 |
| Upper 95% CI of mean | 3.197 | 2.943 | 5.899 | 2.647 |
| Coefficient of Variation | 96.50% | 81.56% | 86.95% | 73.77% |

| Carbohydrate to protein ratio (AU) | | | | |
|---|---|---|---|---|
| | control | CRS | CRSsNP | CRSwNP |
| Number of samples (n) | 4 | 11 | 4 | 7 |
| Minimum | 0.2413 | 0.02271 | 0.07721 | 0.02271 |
| Maximum | 0.7001 | 0.4277 | 0.4277 | 0.1842 |
| Range | 0.4588 | 0.405 | 0.3505 | 0.1615 |
| Mean | 0.3793 | 0.154 | 0.2176 | 0.1115 |
| Std. Deviation | 0.2162 | 0.1209 | 0.168 | 0.06309 |
| Std. Error of Mean | 0.1081 | 0.03823 | 0.08398 | 0.02576 |
| Lower 95% CI of mean | 0.03526 | 0.06748 | -0.04969 | 0.04533 |
| Upper 95% CI of mean | 0.7233 | 0.2404 | 0.4849 | 0.1778 |
| Coefficient of Variation | 57.00% | 78.52% | 77.20% | 56.56% |

| % of total carbohydrates that are fucose | | | | |
|---|---|---|---|---|
| | control | CRS | CRSsNP | CRSwNP |
| Number of samples (n) | 4 | 11 | 4 | 7 |
| Minimum | 0.04222 | 0.01652 | 0.07795 | 0.01652 |
| Maximum | 0.417 | 1.467 | 1.265 | 1.467 |
| Range | 0.3748 | 1.45 | 1.187 | 1.45 |
| Mean | 0.2663 | 0.4547 | 0.5313 | 0.4036 |
| Std. Deviation | 0.1692 | 0.5402 | 0.5448 | 0.5825 |
| Std. Error of Mean | 0.0846 | 0.1708 | 0.2724 | 0.2378 |
| Lower 95% CI of mean | -0.002941 | 0.06824 | -0.3356 | -0.2077 |
| Upper 95% CI of mean | 0.5355 | 0.8411 | 1.398 | 1.015 |
| Coefficient of Variation | 63.54% | 118.8% | 102.5% | 144.3% |

| % of total carbohydrates that are sialic acid | | | | |
|---|---|---|---|---|
| | control | CRS | CRSsNP | CRSwNP |
| Number of samples (n) | 4 | 11 | 4 | 7 |
| Minimum | 2.708 | 6.235 | 6.235 | 9.714 |
| Maximum | 4.639 | 46.06 | 29.09 | 46.06 |
| Range | 1.931 | 39.82 | 22.85 | 36.34 |
| Mean | 3.723 | 20.8 | 18.06 | 22.64 |
| Std. Deviation | 0.8026 | 12.3 | 11.6 | 13.47 |
| Std. Error of Mean | 0.4013 | 3.889 | 5.801 | 5.497 |
| Lower 95% CI of mean | 2.446 | 12.01 | -0.4072 | 8.506 |
| Upper 95% CI of mean | 5.001 | 29.6 | 36.52 | 36.77 |

| | | | | |
|---|---|---|---|---|
| Coefficient of Variation | 21.56% | 59.11% | 64.26% | 59.48% |

| Sialic acid (w/w%) by dry mass | | | | |
|---|---|---|---|---|
| | control | CRS | CRSsNP | CRSwNP |
| Number of samples (n) | 4 | 11 | 4 | 7 |
| Minimum | 0.9739 | 1.252 | 1.252 | 3.632 |
| Maximum | 4.205 | 13.03 | 4.756 | 13.03 |
| Range | 3.231 | 11.78 | 3.504 | 9.401 |
| Mean | 2.025 | 6.132 | 3.539 | 7.613 |
| Std. Deviation | 1.473 | 3.807 | 1.649 | 3.97 |
| Std. Error of Mean | 0.7367 | 1.148 | 0.8247 | 1.5 |
| Lower 95% CI of mean | -0.3198 | 3.574 | 0.9143 | 3.942 |
| Upper 95% CI of mean | 4.369 | 8.689 | 6.164 | 11.28 |
| Coefficient of Variation | 72.77% | 62.09% | 46.61% | 52.14% |

| Fucose (w/w%) by dry mass | | | | |
|---|---|---|---|---|
| | control | CRS | CRSsNP | CRSwNP |
| Number of samples (n) | 4 | 11 | 4 | 7 |
| Minimum | 0.3203 | 0.1216 | 0.3073 | 0.1216 |
| Maximum | 2.309 | 3.249 | 2.129 | 3.249 |
| Range | 1.989 | 3.127 | 1.821 | 3.127 |
| Mean | 1.074 | 1.167 | 1.21 | 1.143 |
| Std. Deviation | 0.8628 | 1.045 | 0.7692 | 1.233 |
| Std. Error of Mean | 0.4314 | 0.3149 | 0.3846 | 0.4661 |
| Lower 95% CI of mean | -0.2985 | 0.4658 | -0.01438 | 0.00296 |
| Upper 95% CI of mean | 2.447 | 1.869 | 2.433 | 2.284 |
| Coefficient of Variation | 80.31% | 89.47% | 63.59% | 107.8% |

| Sodium (w/w%) by hydrated mass | | | | |
|---|---|---|---|---|
| | control | CRS | CRSsNP | CRSwNP |
| Number of samples (n) | 4 | 11 | 4 | 7 |
| Minimum | 0.163 | 0.186 | 0.294 | 0.186 |
| Maximum | 0.273 | 0.337 | 0.337 | 0.319 |
| Range | 0.11 | 0.151 | 0.043 | 0.133 |
| Mean | 0.228 | 0.273 | 0.308 | 0.254 |
| Std. Deviation | 0.0465 | 0.0465 | 0.0197 | 0.0464 |
| Std. Error of Mean | 0.0233 | 0.014 | 0.00984 | 0.0176 |
| Lower 95% CI of mean | 0.154 | 0.242 | 0.277 | 0.211 |
| Upper 95% CI of mean | 0.302 | 0.305 | 0.339 | 0.297 |
| Coefficient of Variation | 20.4% | 17.0% | 6.39% | 18.3% |

| Magnesium (w/w%) by hydrated mass | | | | |
|---|---|---|---|---|
| | control | CRS | CRSsNP | CRSwNP |

| | | | | |
|---|---|---|---|---|
| Number of samples (n) | 4 | 11 | 4 | 7 |
| Minimum | 0.000912 | 0.00117 | 0.00153 | 0.00117 |
| Maximum | 0.00225 | 0.00598 | 0.00598 | 0.00501 |
| Range | 0.00134 | 0.00481 | 0.00445 | 0.00384 |
| Mean | 0.00138 | 0.00297 | 0.00353 | 0.00264 |
| Std. Deviation | 0.000597 | 0.00167 | 0.002 | 0.00151 |
| Std. Error of Mean | 0.000299 | 0.000503 | 0.001 | 0.000571 |
| Lower 95% CI of mean | 0.000433 | 0.00185 | 0.000342 | 0.00124 |
| Upper 95% CI of mean | 0.00233 | 0.00409 | 0.00672 | 0.00404 |
| Coefficient of Variation | 43.2% | 56.2% | 56.8% | 57.2% |

Potassium (w/w%) by hydrated mass

| | control | CRS | CRSsNP | CRSwNP |
|---|---|---|---|---|
| Number of samples (n) | 4 | 11 | 4 | 7 |
| Minimum | 0.0391 | 0.0516 | 0.0638 | 0.0516 |
| Maximum | 0.0749 | 0.177 | 0.177 | 0.117 |
| Range | 0.0358 | 0.125 | 0.113 | 0.0654 |
| Mean | 0.0648 | 0.102 | 0.121 | 0.0916 |
| Std. Deviation | 0.0172 | 0.0352 | 0.0506 | 0.0206 |
| Std. Error of Mean | 0.0086 | 0.0106 | 0.0253 | 0.00778 |
| Lower 95% CI of mean | 0.0374 | 0.0785 | 0.0402 | 0.0725 |
| Upper 95% CI of mean | 0.0922 | 0.126 | 0.201 | 0.111 |
| Coefficient of Variation | 26.5% | 34.4% | 41.9% | 22.5% |

Calcium (w/w%) by hydrated mass

| | control | CRS | CRSsNP | CRSwNP |
|---|---|---|---|---|
| Number of samples (n) | 4 | 11 | 4 | 7 |
| Minimum | 0.00443 | 0.00657 | 0.0106 | 0.00657 |
| Maximum | 0.00805 | 0.0157 | 0.0157 | 0.0153 |
| Range | 0.00362 | 0.00913 | 0.0051 | 0.00873 |
| Mean | 0.00621 | 0.0112 | 0.0127 | 0.0103 |
| Std. Deviation | 0.00148 | 0.00318 | 0.00237 | 0.00342 |
| Std. Error of Mean | 0.000739 | 0.000959 | 0.00118 | 0.00129 |
| Lower 95% CI of mean | 0.00386 | 0.00904 | 0.00891 | 0.00716 |
| Upper 95% CI of mean | 0.00856 | 0.0133 | 0.0164 | 0.0135 |
| Coefficient of Variation | 23.8% | 28.4% | 18.7% | 33.1% |

Elastic Modulus, G' @ 1 Hz (Pa)

| | control | CRS | CRSsNP | CRSwNP | Low hyd. | High hyd. |
|---|---|---|---|---|---|---|
| Number of samples (n) | 4 | 10 | 4 | 6 | 6 | 8 |
| Minimum | 10.37 | 7.675 | 8.841 | 7.675 | 9.834 | 10.37 |
| Maximum | 14.77 | 43.74 | 43.74 | 32.21 | 43.74 | 32.21 |
| Range | 4.396 | 36.06 | 34.89 | 24.54 | 33.9 | 24.54 |

| | | | | | | |
|---|---|---|---|---|---|---|
| Mean | 12.68 | 17.91 | 20.18 | 16.4 | 19.52 | 14.09 |
| Std. Deviation | 1.814 | 11.9 | 16.27 | 9.443 | 12.74 | 8.015 |
| Std. Error of Mean | 0.907 | 3.763 | 8.134 | 3.855 | 5.2 | 2.834 |
| Lower 95% CI of mean | 9.796 | 9.4 | -5.71 | 6.491 | 6.154 | 7.389 |
| Upper 95% CI of mean | 15.57 | 26.42 | 46.06 | 26.31 | 32.89 | 20.79 |
| Coefficient of Variation | 14.30% | 66.43% | 80.63% | 57.58% | 65.25% | 56.88% |

| Viscous Modulus, G” @ 1 Hz (Pa) | | | | | | |
|---|---|---|---|---|---|---|
| | control | CRS | CRSsNP | CRSwNP | Low hyd. | High hyd. |
| Number of samples (n) | 4 | 10 | 4 | 6 | 6 | 8 |
| Minimum | 1.171 | 0.4085 | 1.386 | 0.4085 | 1.462 | 0.4085 |
| Maximum | 2.502 | 13.03 | 13.03 | 6.163 | 13.03 | 2.502 |
| Range | 1.331 | 12.62 | 11.64 | 5.755 | 11.57 | 2.094 |
| Mean | 1.837 | 3.204 | 4.424 | 2.391 | 4.538 | 1.521 |
| Std. Deviation | 0.6214 | 3.797 | 5.74 | 2.05 | 4.512 | 0.6866 |
| Std. Error of Mean | 0.3107 | 1.201 | 2.87 | 0.8368 | 1.842 | 0.2428 |
| Lower 95% CI of mean | 0.848 | 0.4879 | -4.71 | 0.2401 | -0.197 | 0.9466 |
| Upper 95% CI of mean | 2.826 | 5.921 | 13.56 | 4.542 | 9.272 | 2.095 |
| Coefficient of Variation | 33.83% | 118.5% | 129.7% | 85.72% | 99.43% | 45.15% |

| Complex Modulus, G* @ 1 Hz (Pa) | | | | | | |
|---|---|---|---|---|---|---|
| | control | CRS | CRSsNP | CRSwNP | Low hyd. | High hyd. |
| Number of samples (n) | 4 | 10 | 4 | 6 | 6 | 8 |
| Minimum | 10.44 | 7.689 | 9.124 | 7.689 | 9.95 | 7.689 |
| Maximum | 14.99 | 45.64 | 45.64 | 32.33 | 45.64 | 32.33 |
| Range | 4.55 | 37.95 | 36.51 | 24.64 | 35.68 | 24.64 |
| Mean | 12.84 | 18.31 | 20.77 | 16.68 | 20.13 | 14.22 |
| Std. Deviation | 1.871 | 12.36 | 17.1 | 9.568 | 13.43 | 8.012 |
| Std. Error of Mean | 0.9356 | 3.908 | 8.548 | 3.906 | 5.482 | 2.833 |
| Lower 95% CI of mean | 9.867 | 9.474 | -6.431 | 6.636 | 6.035 | 7.521 |
| Upper 95% CI of mean | 15.82 | 27.16 | 47.97 | 26.72 | 34.22 | 20.92 |
| Coefficient of Variation | 14.57% | 67.48% | 82.30% | 57.37% | 66.72% | 56.35% |

| Phase Angle, $\delta$ @ 1 Hz (°) | | | | | | |
|---|---|---|---|---|---|---|
| | control | CRS | CRSsNP | CRSwNP | Low hyd. | High hyd. |
| Number of samples (n) | 4 | 10 | 4 | 6 | 6 | 8 |
| Minimum | 6.271 | 2.778 | 4.549 | 2.778 | 6.412 | 2.778 |
| Maximum | 11.39 | 16.59 | 16.59 | 14.35 | 16.59 | 11.39 |
| Range | 5.124 | 13.81 | 12.04 | 11.57 | 10.18 | 8.617 |
| Mean | 7.934 | 8.297 | 9.246 | 7.664 | 10.73 | 6.288 |
| Std. Deviation | 2.39 | 4.477 | 5.168 | 4.342 | 4.016 | 2.66 |
| Std. Error of Mean | 1.195 | 1.416 | 2.584 | 1.773 | 1.639 | 0.9406 |

| | | | | | | |
|---|---|---|---|---|---|---|
| Lower 95% CI of mean | 4.131 | 5.094 | 1.023 | 3.107 | 6.519 | 4.064 |
| Upper 95% CI of mean | 11.74 | 11.5 | 17.47 | 12.22 | 14.95 | 8.512 |
| Coefficient of Variation | 30.12% | 53.96% | 55.89% | 56.66% | 37.42% | 42.31% |

| Elastic Modulus, G' @ 10 Hz (Pa) | | | | | | |
|---|---|---|---|---|---|---|
| | control | CRS | CRSsNP | CRSwNP | Low hyd. | High hyd. |
| Number of samples (n) | 4 | 10 | 4 | 6 | 6 | 8 |
| Minimum | 259.2 | 234.4 | 252.8 | 234.4 | 251.5 | 234.4 |
| Maximum | 421.9 | 383.2 | 383.2 | 360.8 | 421.9 | 383.2 |
| Range | 162.7 | 148.9 | 130.4 | 126.5 | 170.3 | 148.9 |
| Mean | 310.4 | 299.9 | 307.7 | 294.6 | 301.8 | 303.7 |
| Std. Deviation | 75.49 | 48.46 | 55.9 | 47.65 | 62.59 | 52.03 |
| Std. Error of Mean | 37.74 | 15.32 | 27.95 | 19.45 | 25.55 | 18.4 |
| Lower 95% CI of mean | 190.2 | 265.2 | 218.8 | 244.6 | 236.1 | 260.2 |
| Upper 95% CI of mean | 430.5 | 334.5 | 396.7 | 344.6 | 367.4 | 347.2 |
| Coefficient of Variation | 24.32% | 16.16% | 18.17% | 16.17% | 20.74% | 17.13% |

| Viscous Modulus, G" @ 10 Hz (Pa) | | | | | | |
|---|---|---|---|---|---|---|
| | control | CRS | CRSsNP | CRSwNP | Low hyd. | High hyd. |
| Number of samples (n) | 4 | 10 | 4 | 6 | 6 | 8 |
| Minimum | 130 | 135.3 | 139.1 | 135.3 | 139.1 | 130 |
| Maximum | 240 | 253.5 | 211.4 | 253.5 | 253.5 | 219.2 |
| Range | 110 | 118.2 | 72.22 | 118.2 | 114.4 | 89.2 |
| Mean | 188.3 | 185.5 | 183.7 | 186.7 | 195.9 | 179 |
| Std. Deviation | 50.42 | 34.79 | 33.7 | 38.64 | 45.35 | 32.25 |
| Std. Error of Mean | 25.21 | 11 | 16.85 | 15.78 | 18.51 | 11.4 |
| Lower 95% CI of mean | 108.1 | 160.6 | 130.1 | 146.1 | 148.3 | 152.1 |
| Upper 95% CI of mean | 268.5 | 210.4 | 237.3 | 227.2 | 243.5 | 206 |
| Coefficient of Variation | 26.78% | 18.76% | 18.35% | 20.70% | 23.14% | 18.01% |

| Complex Modulus, G* @ 10 Hz (Pa) | | | | | | |
|---|---|---|---|---|---|---|
| | control | CRS | CRSsNP | CRSwNP | Low hyd. | High hyd. |
| Number of samples (n) | 4 | 10 | 4 | 6 | 6 | 8 |
| Minimum | 329.9 | 313.5 | 320.5 | 313.5 | 321.4 | 313.5 |
| Maximum | 455.6 | 447.5 | 447.5 | 418.9 | 455.6 | 447.5 |
| Range | 125.8 | 134 | 127.1 | 105.4 | 134.3 | 134 |
| Mean | 381.4 | 366.9 | 369.3 | 365.2 | 374.6 | 368.3 |
| Std. Deviation | 53.06 | 43.68 | 54.63 | 40.43 | 45.59 | 47.35 |
| Std. Error of Mean | 26.53 | 13.81 | 27.31 | 16.51 | 18.61 | 16.74 |
| Lower 95% CI of mean | 296.9 | 335.6 | 282.4 | 322.8 | 326.7 | 328.7 |
| Upper 95% CI of mean | 465.8 | 398.1 | 456.3 | 407.6 | 422.4 | 407.9 |
| Coefficient of Variation | 13.91% | 11.91% | 14.79% | 11.07% | 12.17% | 12.85% |

| Phase Angle, δ @ 10 Hz (°) | | | | | | |
|---|---|---|---|---|---|---|
| | control | CRS | CRSsNP | CRSwNP | Low hyd. | High hyd. |
| Number of samples (n) | 4 | 10 | 4 | 6 | 6 | 8 |
| Minimum | 21.21 | 23.24 | 24.01 | 23.24 | 21.21 | 23.24 |
| Maximum | 44.58 | 42.35 | 36.31 | 42.35 | 44.58 | 39.54 |
| Range | 23.37 | 19.11 | 12.29 | 19.11 | 23.37 | 16.3 |
| Mean | 32.45 | 31.89 | 30.96 | 32.51 | 33.76 | 30.76 |
| Std. Deviation | 11.36 | 6.339 | 5.634 | 7.22 | 9.487 | 6.201 |
| Std. Error of Mean | 5.682 | 2.005 | 2.817 | 2.948 | 3.873 | 2.192 |
| Lower 95% CI of mean | 14.37 | 27.35 | 21.99 | 24.93 | 23.81 | 25.58 |
| Upper 95% CI of mean | 50.54 | 36.42 | 39.92 | 40.08 | 43.72 | 35.95 |
| Coefficient of Variation | 35.01% | 19.88% | 18.20% | 22.21% | 28.10% | 20.16% |

### 3. Detailed Rheological Procedures

All rheological measurements were taken on a NETZSCH Kinexus Prime Ultra+ rheometer at 34 ± 0.1°C and with a humidified solvent trap to prevent evaporation. Four independent runs were collected for each sample as described below. Mucus samples were thawed on ice. 27 μL was loaded and sandwiched between a roughened parallel plate geometry (d=8mm) at ambient temperature. Next, the sample was equilibrated for 5 minutes at 34 ± 0.1°C to allow for rebuild after the shear force of pipette transfer. Amplitude sweeps were performed at 1 Hz from 0.01% strain to the end of linear viscoelastic regime (LVER). Collection was stopped when the phase angle increased for three data points. A strain amplitude of 2% was used for all measurements, which was the largest strain in the LVER, and was selected for maximum signal-to-noise in the subsequent frequency sweep. Frequency sweeps were performed from 0.1 to 20 Hz and reversed from 20 to 0.1 Hz at 2%. For each up/down sweep, data for 10 samples/decade were collected. Interpolation between each measured data point was conducted by cubic polynomial at 300 matching frequencies and the up and down sweep values were averaged. Data is exported with 6 points/decade for ease of visualization. For each frequency analyzed (1 and 10 Hz), G', G'', G*, δ were averaged for that sample. Data from four independent runs per sample were collected.

### 4. Detailed Assay Procedures

#### 4.1 Total protein assay detailed procedure

Total protein content was quantified via commercial bicinchoninic acid (BCA) assays (Thermo Scientific). A minimum of three aliquots of each homogenized mucus sample were analyzed in triplicate. Serial dilutions of bovine serum albumin (BSA) standard were prepared. BSA standard or sample (10 μL) was pipetted into a microplate in triplicate. BCA working reagent was added and the microplate was sealed and incubated at 37 °C for 30 min. The microplate was cooled to ambient temperature before taking absorbance readings at 562 nm. A BSA standard curve was used to quantify protein content of the mucus samples.

#### 4.2 Total carbohydrate assay detailed procedure

Total carbohydrate content was quantified via commercial phenol-sulfuric acid assay (Cell Biolabs Inc.) in which concentrated sulfuric acid hydrolyzes complex carbohydrates into monosaccharides and dehydrates them into furfural derivatives that subsequently react with phenol to produce a colorimetric signal. A minimum of two aliquots of each mucus sample were analyzed in triplicate. Serial dilutions of glucose standard were prepared. Glucose standard and homogenized mucus solutions were added to microcentrifuge tubes followed by addition of concentrated sulfuric acid. Samples were incubated at 90 °C for 15 min on a heat block, then cooled at 4 °C. Samples were mixed thoroughly via repeated pipette agitation before transferring 180 μL aliquots of standard or sample to a microplate in triplicate. Background absorbance at 490 nm was collected. To each well, 30 μL of 5% phenol solution was added with a multichannel pipette and reactions were allowed to develop for 5 min at ambient temperature with gentle rocking. Abs 490 was again collected. A glucose standard curve was created to quantify total carbohydrate content of the mucus samples.

#### 4.3 Fucose assay detailed procedure

Fuc content was quantified using a commercial L-fucose dehydrogenase (FDH) assay kit (Megazyme). A minimum of two aliquots of each mucus sample were analyzed in triplicate. Serial dilutions of L-Fuc standard were prepared. Standards or samples (10 μL) were pipetted into a microplate in triplicate. Subsequently, 40 μL of assay buffer and 10 μL of $NADP^+$ were added and the final volume brought to 260 μL with ultrapure water. The plates were gently mixed and incubated for 4 minutes. Background absorbance at 340 nm was collected before the enzymatic reaction. L-FDH enzyme (5 μL) was then added to each well, mixed gently, and incubated at ambient temperature for 10 min. Abs 340 nm was collected every 2 minutes until readings plateaued, indicating reaction completion. An L-Fuc standard curve was created to quantify Fuc content of the mucus samples.

#### 4.4 Sialic acid assay detailed procedure

Sia content was quantified via an improved Warren Method using a commercial kit (Sigma-Aldrich). A minimum of two aliquots of each mucus sample were analyzed in triplicate. Serial dilutions of Sia (*N*-acetylneuraminic acid) standard were prepared. In a microcentrifuge tube, 20 μL of each Sia standard was combined with 5 μL of 10% trichloroacetic acid (TCA). To hydrolyze Sia from the mucus, 20 μL of sample was combined with 40 μL of hydrolysis reagent and 40 μL of ultrapure water at 80 °C for 60 min. Samples were cooled and 25 μL of 10% TCA was added to precipitate peptide/protein in the sample, followed by centrifugation at 14,000 RPM for 10 minutes. Subsequently, 25 μL of sample was transferred to a fresh microcentrifuge tube. Oxidation reagent stock solution was prepared by combining 15 μL hydrolysis reagent, 65 μL oxidation reagent, and 50 μL ultrapure water for each standard and sample. To each sample/standard, 125 μL of oxidation reagent was added and incubated at ambient temperature for 1 hour. Dye reagent (50 μL) was added and samples heated at 95 °C for 12 minutes. Tubes were cooled before adding 100 μL DMSO. Samples were mixed well and centrifuged at 14,000 RPM for 5 minutes. Finally, 250 μL was transferred into individual wells of a microplate and Abs 549 nm were collected. A standard curve was created to quantify Sia content of the mucus samples.
